# Cardiovascular diseases associated with influenza infection: protocol for a systematic review and meta-analysis

**DOI:** 10.1101/2021.08.11.21261941

**Authors:** María del Carmen Caycho Torres, Johnny Clavo Yamahuchi, Cory Ximena Cornejo Ramos, Katherine Ada Diaz Gomez, Oscar Alonso Gayoso Liviac, Omar Fabricio Zanoni Ramos, Victor Vega Zambrano, Cesar Ugarte-Gil

## Abstract

**Background:** Cardiovascular diseases represent important diagnoses that can become fatal if an early and adequate management is not carried out. Since 1930, a possible relationship between these events and influenza virus infection has been proposed.

**Objectives:** To determine the association between cardiovascular diseases and previous infection by influenza virus.

**Materials and methods:** We propose to do a systematic review and meta-analysis according to PRISMA. In order to do so, we will perform an electronic search in the databases of EMBASE, PubMed, Global Index Medicus, Google Scholar and Cochrane Library. Articles will first be selected according to their respective titles and abstracts; and subsequently, the full text of each article will be evaluated. Both phases will be executed by 4 authors. The extracted data will include study details, study methodology and results. The quality of the studies will be verified using standardized tools according to the study design and we will evaluate possible biases. In addition, a meta-analysis of the association measures will be performed using Cochrane’s Software Review Manager 5.4.1 and we will use the GRADE tool to assess the certainty of the results obtained from the analyzed studies.

**Results:** We will publish the results of this systematic review and meta-analysis in a peer-reviewed journal.

**Conclusions:** This systematic review will provide more up-to-date evidence compared to previous work on the association between laboratory-confirmed influenza and cardiovascular diseases.

## BACKGROUND

During the SARS-COV 2 pandemic, many respiratory illnesses caused by other viruses, such as influenza, were neglected. The World Health Organization estimates an approximate 1 000 million infected by the influenza virus to date, with 3 to 5 million causing severe illness and causing an approximate mortality of 300 000 to 500 000 (1). Between 2018 and 2019 in the US, a cumulative incidence of hospitalization for influenza of 65.3 per 100 000 inhabitants was obtained, similar to the periods from 2014 to 2015 and 2016 to 2017. In this, 92.6% of the Hospitalized adults and 55% of hospitalized children had a concomitant medical condition, such as cardiovascular disease, metabolic disorders, obesity, chronic lung disease, asthma, and neurological disorders (2). An important fact is that mortality from cardiovascular diseases in the influenza season increases between 2.3% - 6.3% (3).

Since 1930, a relationship between cardiovascular mortality and influenza infection has been speculated (4). Pathophysiologically speaking, the influenza virus causes endothelial damage and is capable of releasing a cytokine storm, producing tumor necrosis factor (TNF), IL-6 and IL-1β. Furthermore, it is capable of creating a prothrombotic state, since said interleukins increase the expression of plasminogen activator inhibitor 1 (PAI-1) (5).

In a 2015 systematic review, conducted with 16 case-control studies, a significant association was found between recent respiratory infection due to influenza and acute myocardial infarction. In this study, the combined Odds ratio between influenza infection, influenza-like illness (ILI) and respiratory infections were calculated; and it was found that they were more likely to have had these infections in the days prior to an acute myocardial infarction with a combined OR of 2.01 (5).

There are multiple studies that support the association between influenza and cardiovascular events. For example, a study conducted in Houston, Texas collected 1,884,985 patients with acute myocardial infarction, of which 9,885 patients had influenza and 11,485 had other viral infections, a total of 1.1% of the total patients evaluated (6). It was observed that patients with concomitant influenza had a higher rate of hospital fatality, longer hospital stay, development of shock, acute respiratory failure, acute kidney injury and a higher rate of blood transfusion. It was concluded that influenza infection is associated with worse outcomes in patients with acute myocardial infarction. On the other hand, a study carried out in Miami, Florida observed that of 4,285,641 patients with a discharge diagnosis of AMI, 12,830 had concomitant influenza. The patients were between 62 and 85 years old and had more comorbidities than those without influenza infection. Acute myocardial infarction without ST elevation was observed in 90% of cases, while in those without influenza it was 74% (7).

In another case series study in Ontario, Canada, a group of residents older than 35 years in whom infection with different respiratory viruses was found and were hospitalized for acute myocardial infarction in the interval from 2008 to 2015, 364 cases were found of acute myocardial infarction, of which 332 were positive for influenza and only 20 occurred within the risk range (7 days after respiratory infection). Higher risk was found in the first 3 days post infection, in cases of influenza B, in older patients and in patients with first hospitalization for acute myocardial infarction (8).

An association between influenza infection and cardiac arrhythmias has been seen. For example, in a retrospective study carried out in Houston, Texas in 2019, it was seen that there is a significant correlation between high influenza activity and the incidence of ventricular arrhythmias that required shock treatment, in patients with implanted cardiac defibrillators (9). Another study carried out in Taiwan in 2016 indicates that unvaccinated patients with influenza are associated with a significantly higher risk of developing atrial fibrillation (AF), with an Odds ratio of 1.18 (p = 0.03). Whereas, in vaccinated patients, but without infection, the risk of atrial fibrillation was significantly lower with an Odds ratio of 0.88 (p <0.001). Finally, vaccinated patients who had the infection had an Odds ratio similar to patients who were not infected or vaccinated, with an Odds ratio of 1.14 (p = 0.21). It is concluded that influenza infection is significantly associated with the development of AF, with an 18% increase in risk, which can be reduced with vaccination (10).

Additionally, there have been several reports on myocarditis and influenza over the years. The clinical diagnosis of myocarditis, based on symptoms, elevated cardiac enzymes and echocardiography has been reported in 0,4% - 13% of hospitalized patients with documented influenza. (11) There are studies on the 1957 Asian influenza epidemic, where histopathological studies were obtained where it was demonstrated from focal to diffuse myocarditis in 39% of cases. (12) In a Japanese study on the influenza A (H3N2) epidemic between 1998 and 1999, 96 patients with confirmed influenza were evaluated and serum myosin light chain I (MLC-I) was studied as a marker of myocardial damage, of which 11 patients (11.4%) presented elevated MLC-I. (13) In Japan, a contrast was made between the period of the influenza A (H1N1) epidemic from 2009 to 2010 with the following influenza season from 2010 to 2011. It was observed that during the 2009 to 2010 season, the incidence of myocarditis in the pandemic period was greater than during the 2010 to 2011 season, 25 patients versus 4 patients, and the high prevalence of fulminant disease (17/29, 59%) was demonstrated in patients with myocarditis and influenza. (14)

Pericardial involvement in the context of influenza has also been described as a complication, including cardiac tamponade in some case reports (15); however, there is little research in this regard and it may be an underdiagnosed condition.

In the relationship between influenza and pericarditis, there were 2 cases reported in 2016 (16) and 2019 (17); In addition, a case associated with the use of the influenza vaccine was reported in 2018 (18). In a study in Japan, in which 102 participants were evaluated from 2014 to 2017, only 2 cases of pericarditis were reported as a complication due to influenza infection confirmed by nasal swab (19).

The effect of increasing morbidity and complications in patients with heart failure and influenza infection is known. A cohort study showed an increase in hospitalizations for heart failure during the influenza season (20). In a 2020 meta-analysis that involved 6 cohort studies considering 179,158 patients, it was concluded that influenza vaccination was associated with a decrease in mortality in patients with heart failure (21).

The last systematic review that analyzed a possible relationship between influenza infection and cardiovascular effects was in 2015 (5). Little by little, this relationship is becoming more known in the medical community; however, in recent years, recent studies have emerged that require evaluation.

## OBJECTIVES

### Main

- To determine the association between cardiovascular diseases and influenza virus infection.

### Secondary

- To determine the association between acute myocardial infarction and influenza virus infection.
- To determine the association between atrial fibrillation and influenza virus infection.
- To determine the association between pericarditis and influenza virus infection.
- To determine the association between heart failure and influenza virus infection.
- To determine the association between myocarditis and influenza virus infection.

## MATERIALS AND METHODS

### Study design

A systematic review will be carried out according to PRISMA (Preferred Reporting Items for Systematic Review and Meta-Analysis) (22)

### Eligibility Criteria

#### Participants

We will include all patients over 18 years of age infected with any influenza virus serotype who had cardiovascular complications.

#### Exposition

We will consider those patients with diagnosis of influenza by serological tests or by any microbiological test: viral culture, rapid antigen detection test, reverse transcription polymerase chain reaction (RT-PCR), immunofluorescence assays and rapid molecular detection assays.

#### Comparison

Patients with cardiovascular complications as elevated troponins, CPK-MB, pro-BNP, changes in EKG, acute myocardial infarction, cardiac tamponade, heart failure, cardiac arrhythmias.

#### Types of Studies

We will include experimental and observational studies including cross-sectional, case-control, cohorts, randomized and non-randomized clinical trials.

#### Exclusion Criteria

- We will exclude the following publication types: case reports, case series, letter to the editor, editorial, narrative review, cross-sectional studies, duplicated studies, studies without full text and systematic reviews.
- We will exclude pregnant women, pediatric patients, patients without confirmation of influenza or cardiovascular disease.
- We will exclude studies including animals.

#### Literature Search and Data collection

An electronic search will be carried out in the databases EMBASE, PubMed, Global Index Medicus, Google Scholar and Cochrane library. Our search strategy will be using free terms and standardized terms (e.g. MESH), such as Influenza, Cardiovascular diseases, Acute myocardial infarction, Atrial fibrillation, Heart failure, Pericarditis, Myocarditis. There will be no restriction on language or publication date. The search terms are detailed in the supplementary material (table 1).

Articles will be selected and listed in a Microsoft Excel 2019 sheet.

The search results will be exported to the Rayyan web application (23), where duplicates will be eliminated and the studies will be chosen respecting the criteria already described. The application will be used both for the title / abstract selection and for the text revision. The reason why certain studies will be excluded according to the exclusion criteria will be described in the full text phase.

Disagreements will be resolved through discussion until an agreement is reached and the reason for the selection or exclusion of the agreement will be detailed.

The number of studies in each phase will be saved and displayed using a PRISMA flow chart.

#### Data extraction

Data will be extracted from the selected studies located in a Microsoft Excel 2019 spreadsheet (Table 2).

The extracted data will include:

- Study details: First author, country, year of publication.
- Study methodology: Sample size (number of participants in each group / cohort), study design, inclusion criteria and exclusion criteria, influenza diagnostic method, time of presentation of cardiovascular disease with respect to influenza infection, age, gender, follow-up period.
- Result: Measurement of the result method, type of cardiovascular disease, the association estimate (RR, HR or OR) of the result with the 95% CI (confidence interval). The crude and adjusted association estimate will be extracted, as well as the variables to which it adjusts.

## Results

The measure used to present the relationship between our dichotomous results and the EVs will be the hazard ratios (RR) or the odds ratio (OR) with their 95% confidence interval. Our main outcome will be any confirmed cardiovascular damage.

Primary outcome measures:

- Cardiovascular diseases (defined by the authors)

Secondary outcome measures:

- Acute myocardial infarction (defined by the authors)
- Atrial fibrillation (defined by the authors)
- Heart failure (defined by the authors)
- Pericarditis (defined by the authors)
- Myocarditis (defined by the authors)

### Risk of Bias and Methodological quality of Studies

We will assess the risk of bias using different tools according to the study design:

- Cohorts and case-control studies: “National Heart Lung and Blood Institute” (NHLBI) (24)
  ○ According to this assessment tool, the quality of a study can be “good”, “fair” or “poor”. This tool has 14 questions that can be answered as “yes”, “no”, “does not apply”, “cannot be determined” or “not reported”. If any disagreement arises, we will resolve it through discussion. The quality assessment of the studies will be summarized in a table.
- Randomized clinical trials: “Cochrane Risk of bias tool”.
- Non-randomized clinical trials: ROBINS-I

If any discrepancy appears, it will be discussed until a consensus is reached.

### Data synthesis and statistical analysis

We will carry out a narrative synthesis of the evidence found, which will be divided according to our results. A summary of the characteristics of the studies analyzed will be presented using tables.

Statistical analysis will be performed in Cochrane Software Review Manager 5.4.1 (RevMan). The meta-analysis of the association measures (relative risk (RR) or risk ratio (HR) or odds ratio (OR)) will be performed to arrive at a summary estimate. The method for calculating the combined estimate for all outcomes will be the random effects model. We will generate the forest plots based on that, with individual and grouped estimates.

Heterogeneity between studies will be assessed by comparing the methodological characteristics of the included studies, visually looking for signs of heterogeneity in the forest plots. Additionally, it will be evaluated in a quantified way using the I². We will interpret the value of I² following the Cochrane guide (25) with its value:

- 0% to 40%: Non important heterogeneity
- 30% to 60%: moderate heterogeneity
- 50% to 90%: substantial heterogeneity
- 75% to 100%: high heterogeneity

Subgroup analysis will be performed if there is sufficient data. We will analyze the subgroups:

- Gender
- Age
- Patients with acute myocardial infarction
- Patients with atrial fibrillation
- Patients with pericarditis
- Heart failure patients
- Patients with myocarditis

Publication bias will be assessed with the funnel plot if more than ten studies are included. We will consider that there is the possibility of publication bias if there is an asymmetry in the funnel plot of the primary outcome.

We will use the GRADE tool (26) to assess the certainty of the results obtained from the analyzed studies.

## Data Availability

All information used will be from the public domain, thus, there is no need for a ethics committee approval.

## Ethical aspects of the study

No interventions will be performed on humans nor will personal data of included patients be used.

