## Supplementary material for "Cardiovascular diseases associated with influenza infection: protocol for a systematic review and meta-analysis": Table 1

**SUPPLEMENTAL FILES:**

**Table 1:**

| **Database** | **Search strategy:** |
|---|---|
| **Pubmed / Medline** | ("orthomyxoviridae" [MeSH Terms] OR "Influenza" [Title / Abstract] OR "Flu" [Title / Abstract] OR "seasonal flu" [Title / Abstract]) AND ("cardiovascular diseases" [MeSH Terms] OR "cardiovascular diseases "[Title / Abstract] OR (" acute myocardial infarction "[Title / Abstract] OR" heart attack "[Title / Abstract] OR" heart arrest "[Title / Abstract] OR" heart injury "[Title / Abstract] OR "cardiac infarction" [Title / Abstract] OR "cardiopulmonary arrest" [Title / Abstract] OR "coronary infarction" [Title / Abstract] OR "coronary thrombosis" [Title / Abstract]) OR ("atrial fibrillation" [Title / Abstract ] OR "atrial fibrillation" [Title / Abstract] OR "cardiac arrhythmia" [Title / Abstract]) OR ("Stroke"[Title / Abstract] OR "cerebrovascular disorder" [Title / Abstract] OR "cerebrovascular event" [Title / Abstract] OR "cerebrovascular stroke" [Title / Abstract] OR "major stroke" [Title / Abstract] OR "Strokes" [ Title / Abstract])) |
| **EMBASE** | 1. ("orthomyxoviridae" or "Influenza" or "Flu" or "seasonal flu"). Mp. [mp = title, abstract, heading word, drug trade name, original title, device manufacturer, drug manufacturer, device trade name, keyword, floating subheading word, candidate term word]<br><br>2. ("cardiovascular diseases" or "acute myocardial infarction" or "heart attack" or "heart arrest" or "heart injury" or "cardiac infarction" or "cardiopulmonary arrest" or "coronary infarction" or "coronary thrombosis" or " auricular fibrillation "or" atrial fibrillation "or" cardiac arrhythmia "or" Stroke "or" cerebrovascular disorder "or" cerebrovascular event "or" cerebrovascular stroke "or" major stroke "or" Strokes "). mp. [mp = title, abstract, heading word, drug trade name, original title, device manufacturer, drug manufacturer, device trade name, keyword, floating subheading word, candidate term word]<br><br>1 AND 2 |

| | |
|---|---|
| **Global Index Medicus** | 1. (tw: (Influenza, human)) AND (tw: (Cardiovascular Diseases))<br>2. (tw: (Influenza, human)) AND (tw: (Heart Failure))<br>3. (tw: (Influenza, human)) AND (tw: (Myocardial infarction))<br>4. (tw: (Influenza, human)) AND (tw: (Myocarditis))<br>5. (tw: (Influenza, human)) AND (tw: (Pericarditis))<br>6. (tw: (Influenza, human)) AND (tw: (Atrial fibrillation))<br><br>1 AND 2 AND 3 AND 4 AND 5 AND 6 |
| **Cochrane library (CENTRAL)** | #1 MeSH descriptor: [Influenza, Human] explode all trees<br>#2 MeSH descriptor: [Cardiovascular Diseases] explode all trees<br>#3 MeSH descriptor: [Heart Failure] explode all trees<br>#4 MeSH descriptor: [Myocardial Infarction] explode all trees<br>#5 MeSH descriptor: [Pericarditis] explode all trees<br># 6 MeSH descriptor: [Myocarditis] explode all trees<br># 7 MeSH descriptor: [Atrial Fibrillation] explode all trees<br># 8 # 1 AND # 2<br># 9 # 1 AND # 3<br># 10 # 1 AND # 4<br>#eleven # 1 AND # 5<br># 12 # 1 AND # 6<br># 13 # 1 AND # 7 |
| **Google Scholar** | INFLUENZA (HUMAN) AND CARDIOVASCULAR DISEASES<br><br>Selection of the first 200 results |
