## Supplementary material for "Cardiovascular diseases associated with influenza infection: protocol for a systematic review and meta-analysis": Table 2

| | |
|---|---|
| **Study details** | **First author** |
| | **Country** |
| | **Year of publication** |
| **Study methodology** | **Sample size** |
| | **Study design** |
| | **Inclusion criteria** |
| | **Exclusion criteria** |
| | **Diagnostic method** |
| | **Time of presentation of cardiovascular disease with respect to influenza infection** |
| | **Follow-up period** |
| **Outcome** | **Result measurement method** |
| | **Type of cardiovascular disease** |

| | |
|---|---|
| | **The estimate (RR or OR) of the result with 95% CI (confidence interval)** |
